# Perturbations in the mononuclear phagocyte landscape associated with COVID-19 disease severity

**DOI:** 10.1101/2020.08.25.20181404

**Authors:** Egle Kvedaraite, Laura Hertwig, Indranil Sinha, Andrea Ponzetta, Ida Hed Myrberg, Magda Lourda, Majda Dzidic, Mira Akber, Jonas Klingström, Elin Folkesson, Jagadeeswara Rao Muvva, Puran Chen, Susanna Brighenti, Anna Norrby-Teglund, Lars I. Eriksson, Olav Rooyackers, Soo Aleman, Kristoffer Strålin, Hans-Gustaf Ljunggren, Florent Ginhoux, Niklas K. Björkström, Jan-Inge Henter, Mattias Svensson, Karolinska COVID-19 Study Group

**Affiliations:** Center for Infectious Medicine, Department of Medicine Huddinge, Karolinska Institutet, Karolinska University Hospital, Stockholm, Sweden; Childhood Cancer Research Unit, Department of Women’s and Children’s Health, Karolinska Institutet, Stockholm, Sweden; Department of Infectious Diseases, Karolinska University Hospital, Stockholm, Sweden; Department of Physiology and Pharmacology, Section for Anesthesiology and Intensive Care, Karolinska Institutet, Stockholm, Sweden; Function Perioperative Medicine and Intensive Care, Karolinska University Hospital, Stockholm, Sweden; Division of Anesthesiology and Intensive Care, CLINTEC, Karolinska Institutet, Huddinge, Sweden; Division of Infectious Diseases, Department of Medicine Huddinge, Karolinska Institutet, Stockholm, Sweden; Singapore Immunology Network (SIgN), Agency for Science, Technology and Research (A*STAR), BIOPOLIS, Singapore, Singapore; Shanghai Institute of Immunology, Shanghai JiaoTong University School of Medicine, Shanghai, China; Translational Immunology Institute, SingHealth Duke-NUS Academic Medical Centre, Singapore, Singapore; Pediatric Oncology, Theme of Children’s Health, Karolinska University Hospital, Stockholm, Sweden

## Abstract

Monocytes and dendritic cells are crucial mediators of innate and adaptive immune responses during viral infection, but misdirected responses by these cells might contribute to immunopathology. A comprehensive map of the mononuclear phagocyte (MNP) landscape during SARS-CoV-2 infection and concomitant COVID-19 disease is lacking. We performed 25-color flow cytometry-analysis focusing on MNP lineages in SARS-CoV-2 infected patients with moderate and severe COVID-19. While redistribution of monocytes towards intermediate subset and decrease in circulating DCs occurred in response to infection, severe disease associated with appearance of Mo-MDSC-like cells and a higher frequency of pre-DC2. Furthermore, phenotypic alterations in MNPs, and their late precursors, were cell-lineage specific and in select cases associated with severe disease. Finally, unsupervised analysis revealed that the MNP profile, alone, could identify a cluster of COVID-19 non-survivors. This study provides a reference for the MNP response to clinical SARS-CoV-2 infection and unravel myeloid dysregulation associated with severe COVID-19.

## INTRODUCTION

The current COVID-19 pandemic has claimed more than 800.000 lives worldwide during its first 8 months in 2020. Clinical presentation of COVID-19 can vary from asymptomatic to life-threatening acute respiratory distress syndrome and multiple organ failure. Additional potentially lethal complications include profound coagulation abnormalities associated with systemic thrombogenicity combined with a hyperinflammatory state^1,2^. Despite a steadily growing body of information regarding the host immune response to SARS-CoV-2 infection and the pathophysiology behind COVID-19, it is still unclear why certain patients enter the detrimental courses of the disease while others merely present with mild or no symptoms^2^. Thus, there is an urgent need to characterize in depth the immunological and inflammatory aspects of SARS-CoV-2 infection and ensuing COVID-19 disease.

Mononuclear phagocytes (MNPs) in peripheral blood comprise of dendritic cells (DCs) and monocytes, with central roles in orchestrating induction of both innate and adaptive immune responses. Monocytes and DCs are at the front line during an infection, able to recognize, process, and present antigens to immune cells and at the same time produce cytokines and regulate immune responses^3,4^. These cells are divided in subsets with respect to their ontogeny and specialized functions. The monocytes are rapidly recruited to the site of infection, and can be divided into three major subsets; i.e., classical, intermediate, and non-classical^5-7^. Monocytes provide both proinflammatory and/or resolving activities as a supplement to other immune cells present at the site of infection^8,9^. DCs are also a heterogeneous group of cells divided into conventional DCs (cDCs) and type I interferon-(IFN-) producing plasmacytoid DCs (pDCs)^10,11^. Among cDCs, cDC1s are specialized in cross-presenting antigens to CD8+ T cells while cDC2s initiate T helper cell responses, both essential for successful viral clearance^12^. cDC1s constitute a discrete population of cells identified based on highly specific markers such as CLEC9A. cDC2s are more heterogeneous as evident by the functionally distinct CD5+ DC2 and CD5-DC3 subsets, among which inflammatory DC3s, positive for the classically monocyte restricted marker CD14 are found^13-16^. In addition, circulating DC precursors, described as pre-DC^17^ or AS DC^18^, have recently been discovered.

Except for having a central role in the defence against infections, misdirected MNP responses might also contribute to immunopathology. Indeed, the importance of monocytes, monocyte-derived cells and DCs in COVID-19 pathogenesis is emerging^19-27^. Yet, a detailed understanding of developmental and phenotypic alterations, especially with respect to DC lineages and their circulating precursors in COVID-19, is lacking. Moreover, clues on how the altered MNP compartment in COVID-19 is linked to host response to the virus, disease severity, clinical complications, and final outcome are limited. Therefore, we here performed a deep profiling of peripheral blood MNP landscape using 25-color flow cytometry, integrated with publically available single-cell transcriptional data from lung tissue, as well as soluble inflammatory serum factors in SARS-CoV-2 infected patients with moderate and severe COVID-19. Altogether, the present results provide a comprehensive resource of the MNP landscape in response to SARS-CoV-2 infection and ensuing COVID-19 disease.

## RESULTS

### SARS-CoV-2 infection causes declining numbers of circulating cDCs, their progenitors, and pDCs

To study the immune profile of MNPs in SARS-CoV-2 infection, hospitalized patients with ongoing moderate and severe COVID-19 disease were recruited based on strict inclusion and exclusion criteria early in their disease course (Fig. 1A, Supp. Material and Methods, Supp. Table 1, Supp. Table 2). There was no difference in time from onset of symptoms until hospital admission, nor until study sampling, in the two severity groups (Supp. Table 2). An integrative analysis approach was taken where the detailed phenotype of distinct MNP lineages was analyzed in relation to the clinical disease status, combining supervised and unsupervised strategies, with the aim to comprehensively chart the response of circulating MNPs in response to SARS-CoV-2 infection and severity of COVID-19 disease (Fig. 1B). As immune homeostasis is significantly disrupted in COVID-19, canonical lineage markers, such as HLA-DR used to identify discrete subsets of MNPs, are altered in expression^19^. As a point of departure, we designed an MNP-focused 25-color flow cytometry panel (Fig. 1C). After exclusion of granulocytes (CD15+), NK cells (CD7+), ILCs (CD7+), B cells (CD19+), T cells (CD3+), circulating early progenitors (CD34+), basophils (FCER1A+HLA-DR-), and plasma cells (CD38+CD45RA+CD19low), the total MNPs were identified among the cells defined as CD88+ and/or CD116+ (Fig. 1E, D). The MNP identification-strategy allowed clear visualization of DC1, pre-DC and pre-DC2, cDC2, CD5+ DC2, and three subsets of DC3 (CD163-CD14-, CD163-CD14+, CD163+CD14+) (Fig. 2A). This approach was further validated by assessing DC subset-specific markers (Fig. 2B). Next, analysis of absolute numbers of the identified DC subsets revealed that all subsets were decreased in SARS-CoV-2 infected patients (Fig. 2C). In relation to the drastic loss in patients compared to controls, only minor differences where noted between severity groups with pDC and pre-DC loss, if anything, even more profound in the severe COVID-19 patients (Fig. 2D), and with no major differences in-between the different DC sublineages (Fig. 2E). Thus, declining circulating cDCs, their late progenitors, and pDCs is a response feature in SARS-CoV-2 infection.

**FIGURE 1:**
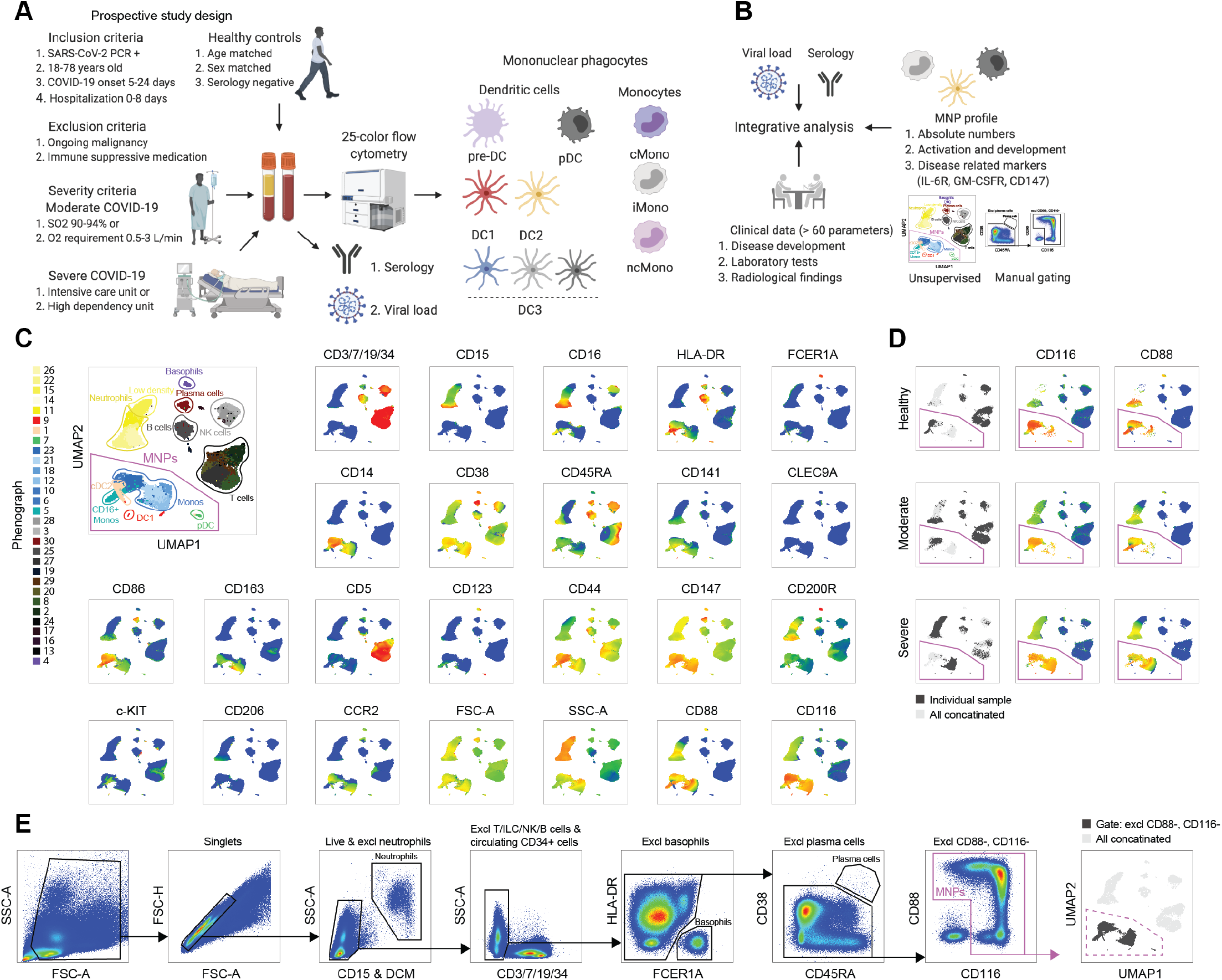
Clinical and experimental study design and analytical pipeline. (A) Experimental design. (B) Analytical pipeline. (C) Flow cytometry data in a UMAP plot with color-coded Phenograph clusters with cell identities established based on the displayed markers; 500.000 live cells from one representative individual from each group (healthy, moderate, severe) is shown after concatenation. (D) Representative individual from each cohort presented in a UMAP; CD116 and CD88 expression highlighted. (E) Gating strategy to identify mononuclear phagocytes (MNPs), after exclusion steps detected in the gate defined as CD88+ and/or CD116+ (i.e. excluding CD88-, CD116 cells), and finally projected back to the UMAP.

**FIGURE 2:**
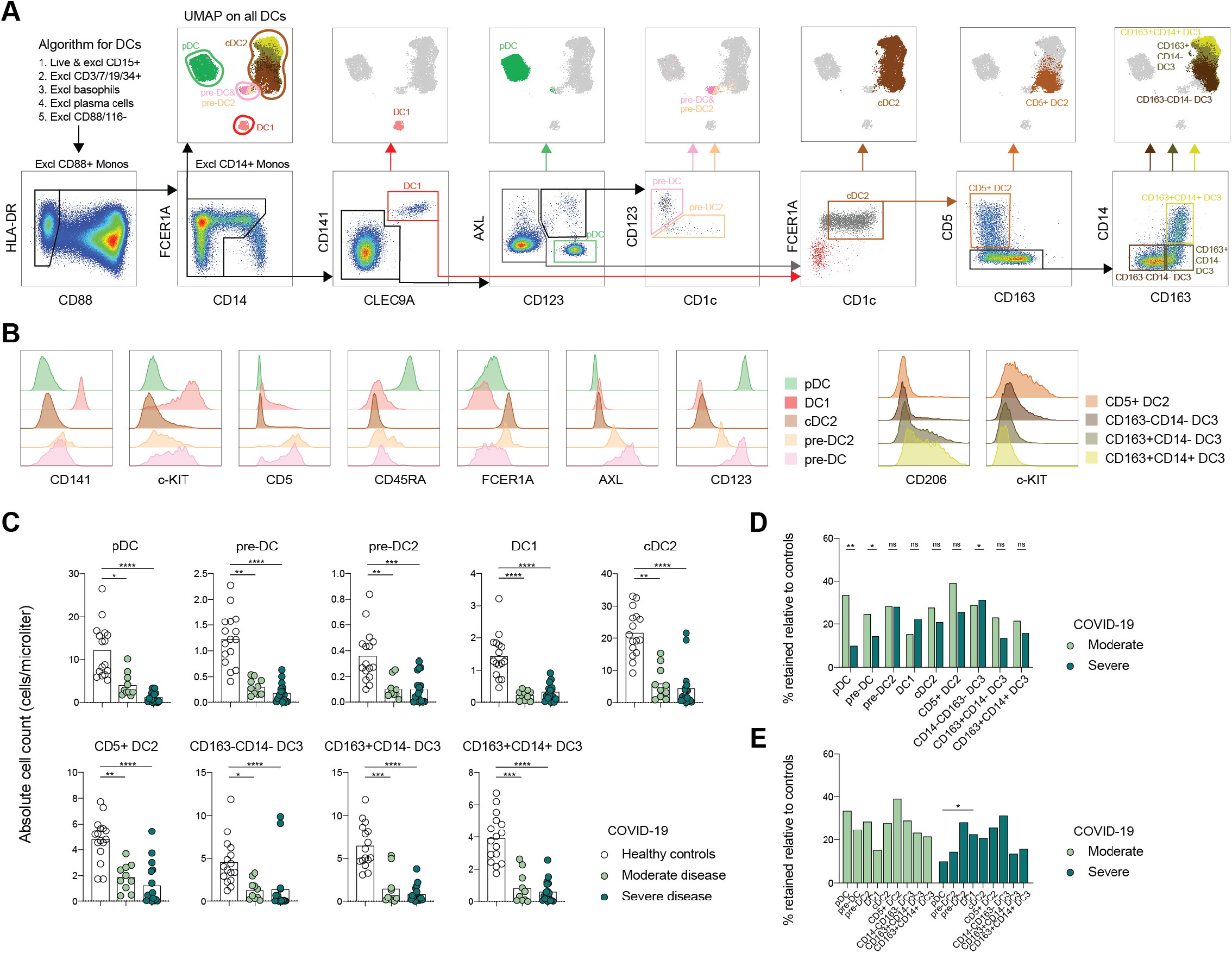
Circulating cDCs, their progenitors, and pDCs decline in numbers in COVID-19 irrespective of disease severity. (A) DC gating strategy (lower panel) projected to the UMAP (upper panel) in all concatenated samples. (B) Key marker expression in the DCs presented across identified DC subsets. (C) Absolute DC numbers in healthy controls, and moderate and severe COVID-19 patients. (D) Absolute DC numbers, each patient related to the mean of the controls as % of maintained cells and compared in moderate vs severe patients for each DC population. (E) % of maintained cells, calculated as described in (D), compared among DC subsets in moderate and severe patients separately. Statistical evaluation using Kruskal-Wallis test and Dunn’s multiple comparisons test (C), Mann-Whitney test (D), and Friedman test (E). P values: *p<0.05, **p<0.01, ***p<0.001, ****p<0.0001.

### DC response to SARS-CoV-2 infection and affected developmental-related phenotypes detected in severe COVID-19

Next, a detailed phenotypic mapping of each DC subset was performed. As a starting-point, Phenograph clustering was performed. The revealed clusters corresponded to all major DC subsets (Fig. 3A). When investigating the phenotype of DC1, upregulated expression of AXL, related to type-I IFN signaling, was a general feature in COVID-19 along with decreased expression of the differentiation marker c-KIT (Fig. 3B-D). Of note, IFN-signaling was also the strongest pathway activated in DC1s at the site of infection as assessed in bronchioalveolar lavage (BAL) samples (Fig. 3E, re-analysis of public scRNA-seq data of BAL immune cells of COVID-19 patients and controls, Supp. Fig. A-R). Circulating pDCs were observed to respond with decreased levels of CD45RA (Fig. 3F-G), while pDCs in BAL were enriched for cytokine and chemokine signaling pathways and further presented with reduced gene expression of *LAMP5*, known to be downregulated in response to TLR9 stimulation and type I IFN signaling^28^, in severe COVID-19 patients (Fig. 3I). Focusing on cDC2s, an increase in frequencies of CD5+ DC2s was detected in moderate COVID-19 (Fig. 3J-K), while a percentage reduction of CD163-CD14-DC3s was noted in severe COVID-19 with an otherwise stable relationship among the DC3 subsets (Fig. 3L). Analyzing the pre-DC subsets (Fig. 3M), a higher percentage of pre-DC2 among the total pre-DC was seen in severe COVID-19 (Fig. 3N). In pre-DCs, an upregulation of the stem cell marker c-KIT was detected in severe COVID-19 (Fig. 3O), and AXL was decreased in response to infection. The levels of AXL correlated positively with serum levels of FMS-like tyrosine kinase 3 ligand (FLT3L), central for DC development (Fig. 3M). Finally, levels of the maturation markers HLA-DR and CD86, the ectoenzyme CD38, the inhibitory receptor CD200R, the GM-CSF receptor (CD116), the IL-6R (CD126), thrombomodulin (CD141), and a possible SARS-CoV-2 spike protein receptor CD147 were assessed across the DC subsets (Fig. 3Q). Higher levels of CD38 were detected specifically in moderate COVID-19 patients, while the maturation markers HLA-DR and CD86 were decreased across all DC subsets (with an exception for DC1s) in severe disease (Fig. 3Q). Finally, in severe disease, all subsets of the cDC2 linage presented with lower levels of CD200R while DC1s specifically downregulated the IL-6R (Fig. 3Q). Taken together, this analysis suggests lineage specific changes and affected developmental phenotype in DC subsets of COVID-19 patients that occur either in response to SARS-CoV-2 infection or specifically in patients with severe COVID-19 disease.

**FIGURE 3:**
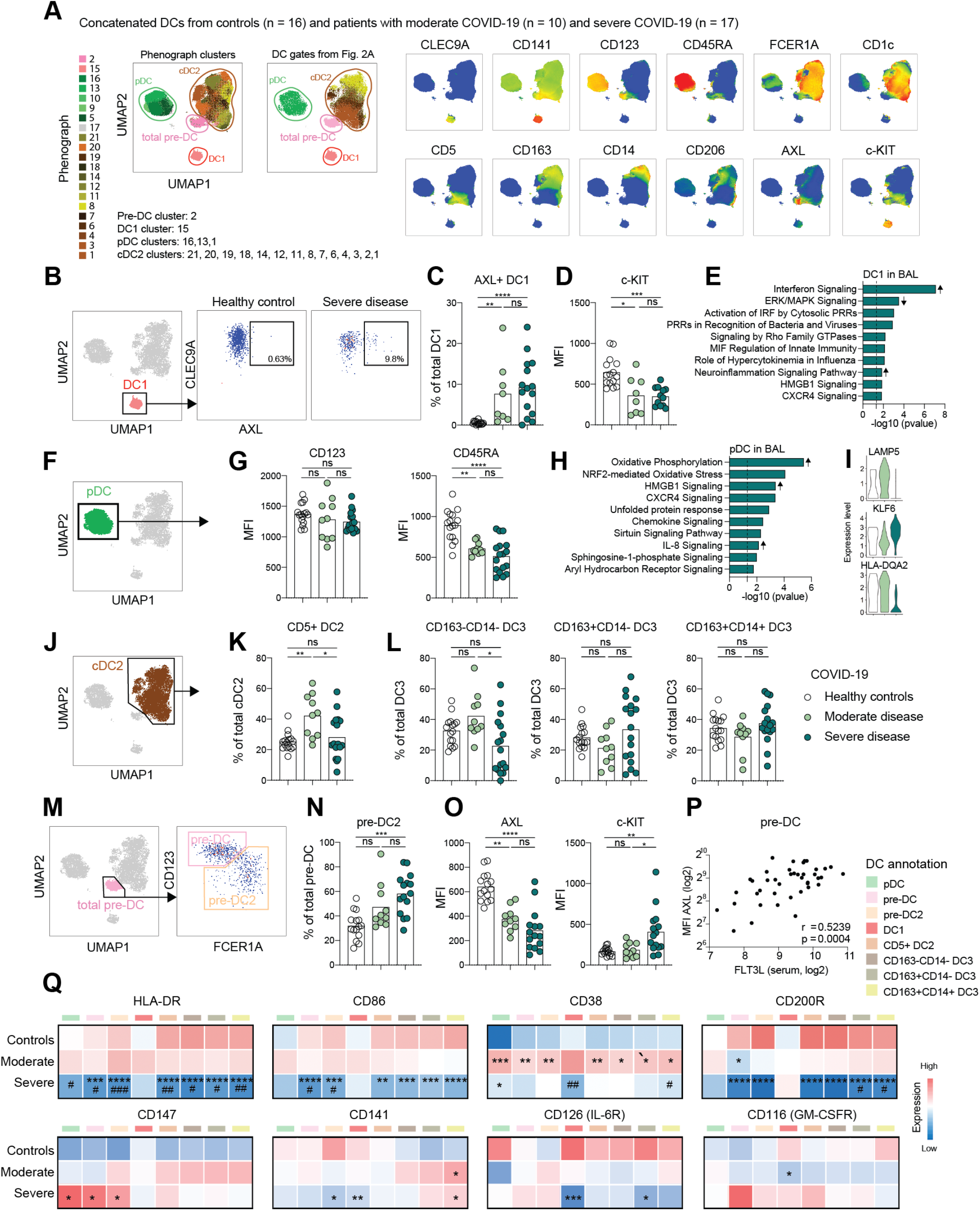
Altered activation and developmental phenotype of DCs in COVID-19. (A) All DCs from patients and controls concatenated in a UMAP, as described in Fig. 2A, with color-coded Phenograph clusters and manual DC gates; cell identity was established based on marker expression. (B) DC1s gated in a UMAP and subjected to analysis of % of AXL+ cells, representative healthy control and severe COVID-19 patient shown in a FACS plot. (C) Quantification of the % of AXL+ DC1s in the three cohorts. (D) MFI of c-KIT quantified in total DC1s, gated in DC UMAP as shown in (B). (E) Ingenuity Pathway Analysis of DEGs upregulated in DC1s in BAL of severe COVID-19 patients compared to healthy controls; arrow indicate direction of upregulation based on Z-score; data re-analyzed from previously published report ^22^, as described in “Single cell analysis and correlation plots”. (F) pDCs gated in a UMAP for downstream analysis. (G) MFI of CD123 and CD45RA quantified in total pDCs in the three cohorts. (H) Ingenuity Pathway Analysis of DEGs upregulated in pDCs in severe patients compared to moderate COVID-19; arrow indicate direction of upregulation based on Z-score; data re-analyzed from previously published report ^22^, as described in “Single cell analysis and correlation plots”. (I) Genes in pDCs differentially expressed between severe and moderate COVID-19 patients, presented in the three cohorts; data re-analyzed from previously published report ^22^, as described in “Single cell analysis and correlation plots”. (J) cDC2s gated in a UMAP for downstream analysis. (K) Quantification of the % of CD5+ DC2 among total cDC2s in the three cohorts. (L) Quantification of the % of the three DC3 subsets among total DC3s (as presented in Fig. 2A, DC3 subsets were defined from the cDC2 gate in (J)) in the three cohorts. (M) Total pre-DCs gated in a UMAP subdivided in pre-DCs and pre-DC2s. (N)Quantification % pre-DC2 in total pre-DCs. (O) MFI of AXL and c-KIT in total pre-DCs in the three cohorts. (P) Correlation between AXL MFI in pre-DCs and soluble FLT3L. (Q) Heatmap showing marker expression in the three cohorts, across the DC subsets, only samples with more than 10 cells included. Statistical evaluation was made separately in each indicated subset by comparing the MFI in the three cohorts; significance for healthy to moderate and healthy to severe comparisons is indicated by * and for moderate to severe comparison is indicated by #. Statistical evaluation using Spearman test for correlation, Kruskal-Wallis test and Dunn’s multiple comparisons test for all other analysis. Samples with less than 10 cells in any of the DC subsets annotated in (Q) were excluded from the analyses. Significance level: *p<0.05, **p<0.01, ***p<0.001, ****p<0.0001.

### Major phenotypic alterations within monocyte subpopulations of COVID-19 patients

Next, we focused on monocytes and their subsets (Fig. 4A). As expected, three major populations of monocytes were identified based on CD14 and CD16 expression; i.e., CD14+CD16- cMonos, CD14+CD16+ iMonos, and CD14lowCD16++ ncMonos (Fig. 4A). UMAP-analysis verified the present gating strategy and demonstrated separation between monocytes and DCs (Fig. 4A). This was also the case for DC3s, which in many aspects are similar to monocytes in phenotype, especially due to their expression of CD14 (Fig. 4A). In accordance with previous publications, iMonos expressed the highest levels of HLA-DR while cMonos displayed relatively high levels of CCR2 (Fig. 4B). Assessment of absolute counts of monocyte populations in COVID-19 patients and controls revealed no change in cMono numbers, significantly increased iMono numbers in response to infection, especially in moderate patients, and declining numbers of ncMonos in all patients (Fig. 4C). A similar pattern was observed when monocyte subset frequencies were assessed (Fig. 4D). While increased CD38 expression across monocyte subsets was a general feature unrelated to disease severity, higher levels of CCR2 on ncMonos and iMonos, as well as lower levels of HLA-DR and CD86 with increased expression of CD163 in all monocyte subsets were found in severe COVID-19 (Fig. 4E). Furthermore, thrombomodulin (CD141) expression was increased in severe COVID-19 in ncMonos and iMonos while it was also increased in moderate patients in cMonos (Fig. 4E). In summary, this shows redistribution and an immature phenotype of the monocyte compartment in COVID-19 that is linked to disease severity.

**FIGURE 4:**
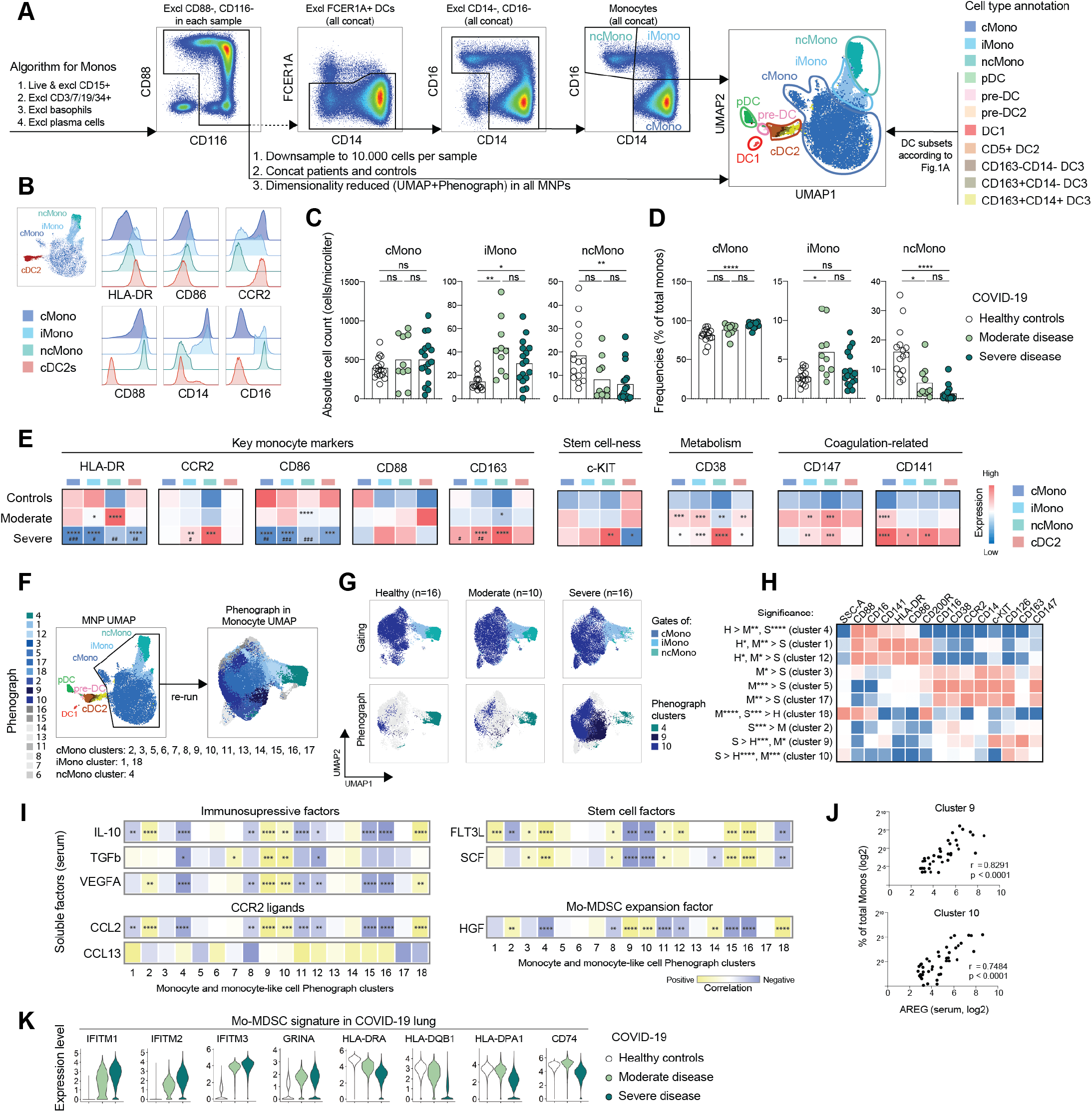
Major phenotypic alterations within monocytes and monocyte-like cells in COVID-19 patients. (A) Monocyte gating strategy projected together with DCs, gated as described in Fig. 2A, to the MNP UMAP. (B) Key monocyte markers in monocyte subsets, compared to cDC2s, in all concatenated samples. (C) Absolute numbers of monocytes in the three cohorts. (D) Monocyte frequencies in the three cohorts. (E) Heatmap showing marker expression in the three cohorts across the monocyte subsets with cDC2s included as a reference. Statistical evaluation was made separately in each indicated subset by comparing the MFI in the three cohorts; significance for healthy to moderate and healthy to severe comparisons are indicated by * and for moderate to severe comparison by #. (F) MNPs from patients and controls concatenated in a UMAP with color-coded manually gated cell subsets (left), rerun on monocytes presented in a UMAP with color-coded Phenograph clusters (right). (G) Color-coded manual gates (upper panel) and indicated Phenograph clusters (lower panel) in monocyte UMAP of concatenated samples. (H) Expression of markers in selected significantly differential Phenograph clusters among the three cohorts (see also Supp. Fig. 2). (I) Heatmap of correlation between Phenograph clusters 1-18 and soluble factors in serum. (J) Correlation between soluble serum AREG and % of Phenograph clusters 9 and 10 in total monocytes. (K) MDSC signature genes in lung MNP compartment differentially expressed between severe COVID-19 patients and controls, presented in the three cohorts; data re-analyzed from previously published report ^22^, as described in “Single cell analysis and correlation plots”. Statistical evaluation using Kruskal-Wallis test and Dunn’s multiple comparisons test for comparison between the three cohorts and Spearman test for correlations, ‘bimod’ test for differentially expressed genes. Significance level: *p<0.05, **p<0.01, ***p<0.001, ****p<0.0001

### Dimensionality reduction reveals MDSC phenotype in severe COVID-19

To analyze the global monocyte landscape in an unbiased manner and integrate the contribution of all tested markers, we next re-clustered all monocytes revealing high heterogeneity of cells that fell into the cMono category (Fig. 4F). Indeed, cMonos stratified into 15 Phenograph clusters whereas only one or two clusters were found for iMono and ncMono (Fig. 4F, Supp. Fig. 1A). When assessing these clusters in relation to disease severity, only one cluster (#4, corresponding to ncMonos) was reduced in all patients compared to controls whereas clusters 9 and 10 were specifically increased in severe compared to moderate patients and healthy controls (Fig. 4G, Supp. Fig. 1A). Next, when clusters with differential presence among the three groups (Supp. Fig. 1A) were ordered sequentially (controls, moderate and severe patients), a distinct phenotypic pattern emerged (Fig. 4H). In clusters enriched in moderate and severe patients, falling into the cMono category, higher expression of the GM-CSF receptor (CD116), IL-6R (CD126), CD147, CD38, and CCR2 was found. Monocyte clusters specific to severe COVID-19 patients (clusters 9, 10) compared to the ones from patients with moderate disease (clusters 3, 5, 17) had lower levels of HLA-DR and CD86. This phenotype was reminiscent of a monocytic myeloid derived suppressor cell (Mo-MDSC)-like phenotype^29,30^. In addition, the Mo-MDSC-like cluster 9 expressed high levels of c-KIT and CD163 (Fig. 4H).

Furthermore, a positive correlation was found between cMono Phenograph clusters specific to severe COVID-19 disease (clusters 2, 9, 10, 18) and the soluble immunosuppressive factors IL-10, TGF-β, and VEGFA as well as with AREG (Fig. 4I, J), known to be involved in tolerance and tissue repair^31^. In contrast, these clusters showed a negative association with stem cell factors important for myeloid cell development and differentiation (Fig. 4I). The Mo-MDSC-like clusters also correlated positively with hepatocyte growth factor (HGF), known to support Mo-MDSC expansion^32^ (Fig. 4I). To search for evidence of an Mo-MDSC signature at the site of infection, publicly available single cell RNA-seq data of BAL from healthy controls, moderate, and severe COVID-19 patients was used^22^ (Supp. Fig. 2A-P). The expression of Mo-MDSC signature genes, comparing monocyte versus Mo-MDSC^33^, was significantly higher in MNPs from severe patients compared to controls (e.g. *IFITM1*, *IFITM2*, *GRINA*, *SOCS3*, and *CD84*) (Fig. 4K, Supp. list 1). In line with the specific immune suppressive Mo-MDSC gene expression pattern, MHC class II molecules and *CD74* were found at lower levels in severe COVID-19 patients compared to controls (Fig. 4K). Altogether, this analysis revealed monocyte heterogeneity related to COVID-19 disease severity and an expansion of monocytes with a MDSC-like phenotype only in severe COVID-19 patients.

### Viremia and seroconversion are associated with recovery of inflammatory DC3s

The patients all presented with elevated inflammatory markers but clinical hyperinflammation and signs of coagulation disturbances were most pronounced in the severe COVID-19 patients (Fig. 5A, Supp. cohort description). Integrative correlation mapping of clinical parameters, including peak measurements recorded during hospitalization up until study sampling (Fig. 5B) and those taken within 24 hours from MNP profiling (Supp. Fig. 3A) revealed that levels of inflammatory cytokines and LDH clustered together with fatal outcome and viremia. Indeed, subgroups of moderate and severe COVID-19 patients had detectable virus in serum (serum PCR+, measured at study sampling), while the majority of seroconverted (SARS-CoV-2 IgG+, measured at study sampling) patients were critically ill (74%, 14/19) (Fig. 5C). Significantly higher numbers of pre-DC2 as well as of inflammatory DC3 (CD163+CD14+ DC3 and CD163+CD14- DC3) were found in patients without ongoing viremia (serum PCR-) and the same DC3 populations were elevated in seroconverted patients (Fig. 5C, Supp. Fig. 3B). Following a similar trend, ncMonos were also present at higher numbers in seroconverted patients (Fig. 5C, Supp. Fig. 3B). As expected, patients that had seroconverted (IgG+) were sampled slightly later in their disease course (Fig. 5D). Absolute numbers of the inflammatory DC3 subset (CD163+CD14- DC3) as well as iMonos and cMonos increased over time after onset of symptoms (Fig. 5E, Supp. Fig. 3C), suggesting that seroconverted patients had passed the time-point when their DC3s were at their lowest levels. Thus, subset-specific changes in MNPs absolute numbers depended on the time from symptom onset, and were associated with a time-dependent recovery of DC3 and increase of monocytes and monocyte-like cells.

**FIGURE 5:**
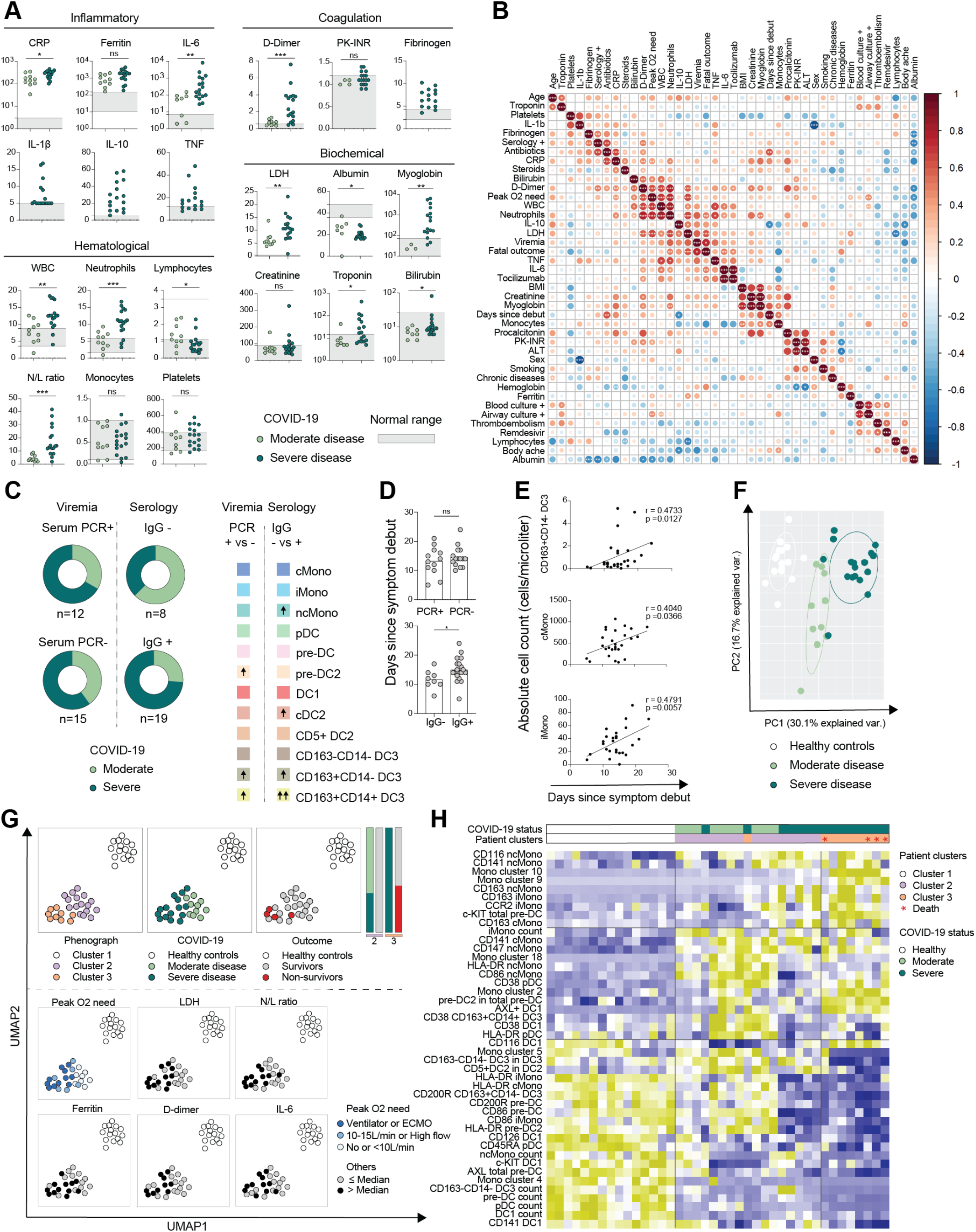
Clinical hyperinflammation, the MNP landscape, and COVID-19 outcome. (A) Levels of clinical analytes in moderate and severe COVID-19 patients, reference levels indicated as a grey shaded area; CRP, D-dimer in mg/L; procalcitonin, ferritin, myoglobin in μg/L; hemoglobin, albumin in g/L; WBC, neutrophils, lymphocytes, monocytes, platelets as 10^9^/L; troponin in ng/L; fibrinogen in g/L; creatinine, bilirubin in μmol/L; LDH in U/L; IL-6, IL-1β, TNF, IL-10 in ng/L. (B) Integrated correlation clustering map of clinical parameters; the color of the circles indicated positive (red) and negative (blue) correlations, color intensity represented correlation strength as measured by the Pearson’s correlation coefficient. (C) Distribution of moderate and severe COVID-19 patients with respect to viremia (SARS-CoV-2 PCR+/-, measured at sampling) and serology (SARS-CoV-2 IgG- /+, measured at sampling) shown in donut charts (left); differences in absolute numbers of MNP populations between SARS-CoV-2 PCR+ and SARS-CoV-2 PCR- patients, and between SARS-CoV-2 IgG- and SARS-CoV-2 IgG+ patients (right), upward arrows indicate significant increase (see also Supp. Fig. 3B). (D) Differences in days since symptom debut until sample collection in SARS-CoV-2 PCR+/- and SARS-CoV-2 IgG-/+ patients. (E) Correlation between days since symptom debut until the sample collection and absolute numbers of cells within MNP populations (see also Supp. Fig. 3C). (F) Principal component analysis of 108 MNP parameters in the three cohorts (see also Supp. Fig. 3D). (G) Dimensionality reduction of 108 MNP parameters for each patient or control presented in UMAP color-coded annotated by Phenograph clusters, disease status, and outcome (above dashed line), as well as levels of clinical parameters, i.e. peak O2 need, LDH, neutrophil/lymphocyte (N/L) ratio, D-dimer, ferritin, and IL-6, converted to categorical variables based on a median value (below dashed line). (H) Hierarchical clustering of patients from the three Phenograph clusters and 44 selected pre-clinical parameters. Statistical evaluation using Mann-Whitney for comparison between the two groups, Spearman test for correlations of non-normally distributed data and Pearson test for correlations normally distributed data. Significance level: *p<0.05, **p<0.01, ***p<0.001, ****p<0.0001.

### Independent assessment of the MNP landscape identifies clinical subgroups and disease outcome-correlates in COVID-19

Finally, to get an overall view of the MNP landscape in relation to COVID-19 disease severity, we performed a principal component analysis of the 108 MNP parameters measured. Here, COVID-19 patients clearly separated from healthy controls and within the patient groups, moderate and severe additionally clustered apart (Fig. 5F). The MNP parameters that contributed highly to separate the groups were: higher DC counts and Mono cluster 4 (non-classical monocytes), higher c-KIT expression in DC1, higher CD45RA expression in pDCs, and higher HLA-DR, CD86 and CD200R expression in cDC2 lineage in controls; higher CD38 expression in MNPs, higher frequency of CD5+ DC2s among cDC2s in moderate patients COVID-19; and an expansion of Mo-MDSC-like clusters (9 and 10), higher c-KIT levels in DC progenitors, and higher frequencies of pre-DC2s in severe patients (Fig. 5F, Supp. Fig. 3D). Dimensionality reduction using UMAP and Phenograph was performed using the same MNP variables, revealing 3 major clusters, where cluster 1 corresponded to healthy controls, cluster 2 corresponded to all moderate and some severe COVID-19 patients, and cluster 3 corresponded to severe COVID-19 patients only (Fig. 5G). All patients with fatal outcome were found in cluster 3 and these further had a high oxygen requirement, showed higher levels of LDH (peak measurement), high neutrophil/lymphocyte ratio, and elevated ferritin (measured within 24 hours from MNP profiling) (Fig. 5G, Supp. Fig. 3E). To visualize the key MNP profile components specific for each cluster, 44 of the most significant MNP parameters analyzed so far were selected and subjected to hierarchical clustering. Monocyte clusters 9 and 10, corresponding to Mo-MDSC-like cells, were specific to patient cluster 3 with the non-survivors. These parameters also clustered together with CD163 expression on all monocyte subsets, CCR2 expression on iMonos, and c-KIT in total pre-DCs. For moderate disease, higher expression of CD38 in all DC lineages, CD141 on cMono, and higher levels of monocyte cluster 18 (iMono, the only cluster expressing high levels of CD200R among the patient specific clusters), were the determining parameters. A set of markers were lost in severe patients including HLA-DR and CD86 in both monocytes and DCs, and CD200R in DC subsets (Fig. 5H). Moreover, the CD200R decrease in non-survivors was restricted to the cDC2 lineage, with the most significant differences seen in progenitors (pre-DCs and pre-DC2) (Supp. Fig. 3F). Finally, to address if this was entirely dependent on severity status rather than outcome, CD200R levels were assessed within severe patients only. A similar pattern was observed, suggesting that CD200R is diminished in the cDC2 lineage and is related not only to disease severity but also to fatal outcome (Supp. Fig. 3F). Altogether our results suggest lineage restricted phenotypic and developmental shifts in MNPs and their late precursors and show that MNPs, alone, could identify a cluster of COVID-19 non-survivors (Fig. 5 and 6).

## DISCUSSION

While broad efforts towards getting an overview of immune cell- and soluble factor alterations in COVID-19 are under way, a deep and comprehensive understanding of the mononuclear phagocyte (MNP) system, including circulating progenitors, is lacking. Here, we used high-dimensional flow cytometry to characterize the MNP landscape in well-defined prospective cohorts of SARS-CoV-2 infected patients with moderate and severe COVID-19 disease as well as matched SARS-CoV-2 IgG negative healthy controls. This allowed a robust mapping of phenotypic and developmental alterations in circulating DCs and monocytes that occurred either in response to clinical infection or specifically in severely sick patients (Fig. 6). By combining conventional gating with unsupervised dimensionality reduction, we provide a reference for absolute numbers and lineage dependent phenotypic shifts in MNP subsets in response to SARS-CoV-2 and reveal an affected developmental phenotype in the cDC2 lineage and expansion of monocytic MDSC-like cells in critically ill patients. In addition, the results were related to data from the site of infection and severe inflammatory impact by means of reanalysing BAL scRNA-seq data and relevant soluble factors in circulation. Finally, and strikingly, alterations within the MNP landscape, alone, were found to separate healthy controls from patients and further identify distinct patient clusters such as moderate and severe as well as within severe patients even those with fatal outcome. Thus, the present high-dimensional mapping of the MNP landscape yields new insights into the MNP response during SARS-CoV-2 infection and in relation to COVID-19 disease severity and provides a comprehensive framework for future studies (Fig. 6).

**FIGURE 6:**
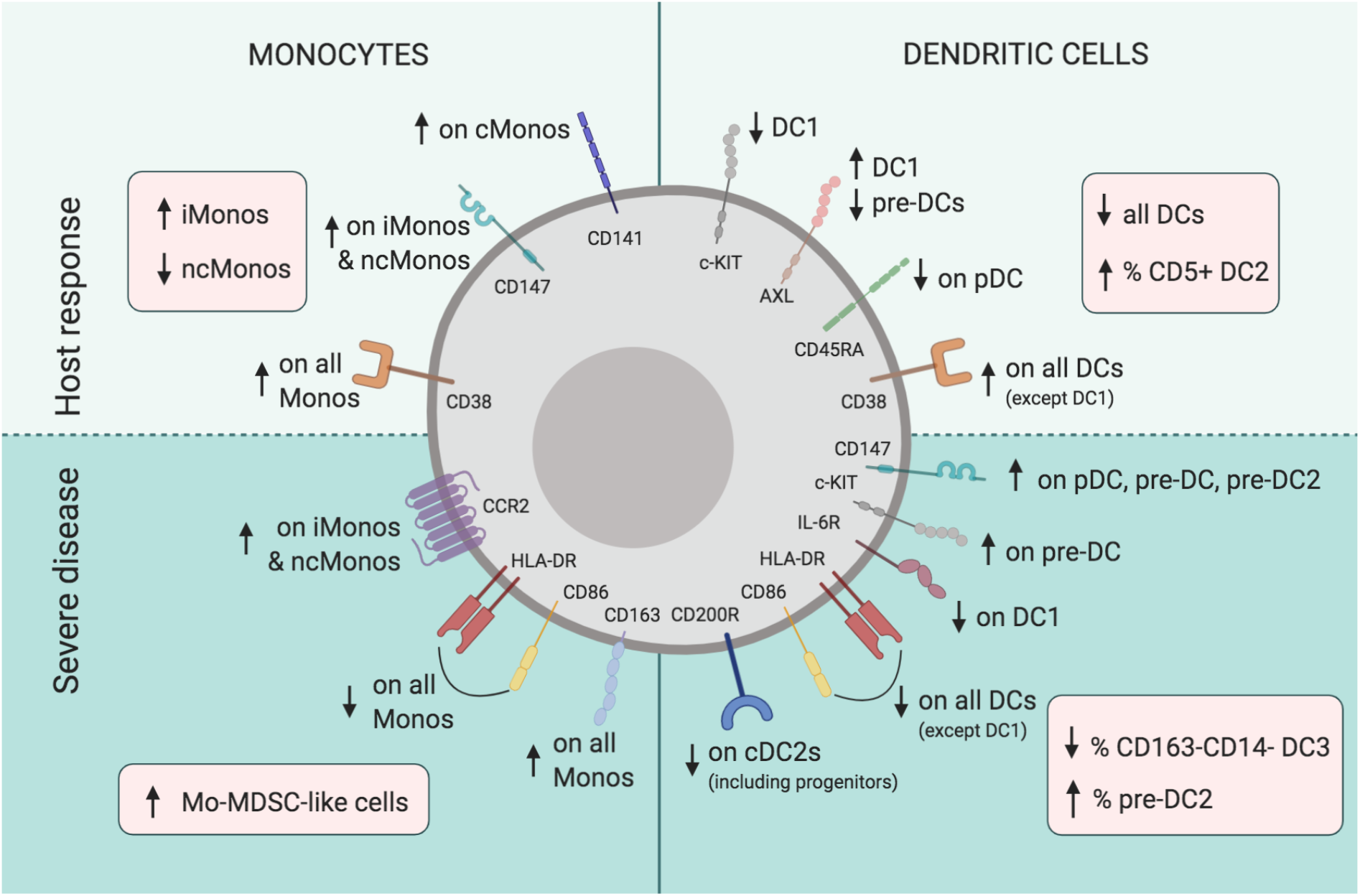
Summary of MNP response to SARS-CoV-2 and in relation to COVID-19 disease severity. Monocytes (left side) and dendritic cells (right side), respond to SARS-CoV-2 (upper panel) with decreased circulating levels of all DC subsets, but higher CD5+ DC2 frequency, activated phenotype (upregulation of ecto-enzyme CD38, expansion of CD14+CD16+ iMonos, decrease of CD14lowCD16++ ncMonos, and lower CD45RA levels on pDCs), and features pointing to viral sensing and type I IFN imprint found in DC1s (e.g. lower levels of c-KIT, expansion of AXL+DC1s). Severe COVID-19 (lower panel) is associated with altered developmental DC phenotype (e.g. increased frequencies of late pre-DC2 progenitors and increased stem cell marker c-KIT expression in pre-DCs, as well as immature phenotype on all DC subsets with lower HLA-DR and CD86), expansion of Mo- MDSC-like cells and decrease in inhibitory receptor CD200R, specific to cDC2 lineage. Arrows in red boxes indicate changes in counts and frequencies of cell populations. Arrows in green areas illustrate altered expression of receptors within indicated populations.

Knowledge on alterations of specific DC subsets and their circulating progenitors in acute viral infection in humans is limited, but the depletion of all DCs and their precursors detected here is in line with previous studies showing DC disappearance in circulation during acute viral insults^20,34,35^. Given their central role in antigen recognition and control of adaptive immunity^36^ as well as their presence in COVID-19 lung^22^, it is plausible that DC disappearance from the circulation is related to their influx to tissues and lymph nodes to lead and assist initiation of adaptive immune responses aiming at virus elimination. In support of this, we find recovery of absolute numbers of pre-DC2s as well as subsets of DC3s in patients where virus has been cleared from circulation and seroconversion occurred. Other features of host response included decreased levels of c-KIT in DC1, expansion of AXL+DC1 and CD5+ DC2s, similarly to monocytes increased expression of the ecto-enzyme CD38 in most DC subsets, and decreased CD45RA on pDC possibly indicating activation as also detected in lung tissue. It has been shown that DC1s downregulate c-KIT upon viral stimulation in an IFN-dependent manner^37^, and it is thus possible that lower c-KIT levels in DC1 found here indicate DC1 SARS-CoV-2 sensing. In support of this, pathways analysis of DC1s in COVID-19 lung revealed PRR-related pathways and an IFN-response imprint. The expansion of AXL+ DC1 in circulation could further support an IFN-affected phenotype, as AXL has been shown to be upregulated on myeloid cells upon type I IFN stimulation^38^.

In severe COVID-19 disease, the DC phenotype showed affected maturation status with lower levels of HLA-DR and CD86 in all DC subsets, with an exception for DC1, and an altered developmental phenotype with expansion of late progenitors pre-DC2 and increased expression of the stem cell marker c-KIT in pre-DCs. Additional lineage specific DC alterations in severe COVID-19 included downregulation of IL-6R in DC1 and a drastic decrease of the inhibitory receptor CD200R in the cDC2 lineage, including circulating progenitors. Decreased CD200R levels might be of particular interest in development of severe COVID-19, given that CD200R signalling is known to modulate immune responses to pathogenic stimuli, control myeloid cell function, inhibit proinflammatory cytokine expression, and inhibit tissue damage caused by myeloid-derived cells^39,40^. Moreover, it has been suggested that CD200R signalling plays an important role in T cell priming during viral infection^41^, but remains to be studied in further detail in COVID19. Our findings suggest that in severe COVID-19, DCs, and in particular the cDC2 lineage, are immature with alterations in the developmental phenotype, lack upregulation of the activation marker CD38 that is seen in moderate patients, and loose the inhibitory receptor CD200R, all possibly contributing to inefficient regulation of pro-inflammatory conditions in severe COVID-19.

In line with previous reports, we observed an expansion of intermediate CD14+CD16+ monocytes in COVID-19 patients^21,42^. This appeared to be a general feature of clinical SARS-CoV-2 infection. In addition, all monocyte subsets responded with increased CD38 expression, that in contrast to DCs, was independent on disease severity. It is plausible that iMonos, exhibiting a strong IFN-signature in COVID-19^27^, contribute to successful viral clearance in moderate COVID-19 patients. In addition, the Phenograph cluster we identified that corresponded to iMonos (cluster 18, Fig. 4H) had high levels of the inhibitory receptor CD200R, supporting the hypothesis that robust innate antiviral responses as well as efficient control of the pro-inflammatory state are important in order to have a moderate course of COVID-19^43^. On the other hand, in line with previous observations^19,27,44^, monocytic cell clusters with an immature phenotype (in particular cluster 9 and 10), identified as Mo-MDSC-like cells, were linked to severe disease and a patient cluster that included all non-survivors. Further studies are required to understand whether Mo-MDSC-like cells described here in fact have suppressor functions, e.g., through a supply of immunosuppressive cytokines, that in the present study showed a positive correlation with Mo-MDSC-like cell frequencies. It is possible that a myeloid differentiation bias in the bone marrow, reflected by the expansion of immature Mo-MDSC-like cells and the lack of adequate expansion of CD14+CD16+ iMonos, the latter being better equipped to clear virus through the IFN-response, contribute to the immunopathogenesis of COVID-19. In support of this, we found a negative correlation between Mo-MDSC and the growth factors FLT3L and SCF, which are required for myeloid differentiation. This coincided with a positive correlation between Mo-MDSC and HGF, known to mediate Mo-MDSC expansion^32^ (Fig. 5I). This would fit with the model of sequential transition, suggesting that a certain proportion of cMonos eventually become iMonos, followed by their development into ncMonos^45^. While the elevated CCR2 on ncMonos found here may present an explanation for their more pronounced disappearance from the circulation in severe COVID-19, a sequential transition model might also explain the decrease of ncMonos detected in COVID-19 patients by us (Fig. 4C, D) and others^20,27,35,42^.

Hyperinflammation in COVID-19 is characterized by high systemic cytokine levels combined with coagulation abnormalities and thromboembolism^46-48^, also indicated in our study by increased levels of fibrinogen and D-dimer. Increased levels of thrombomodulin (CD141), a cofactor for thrombin reducing blood coagulation, possibly possessing an anticoagulant potential, were observed on cMonos in both moderate and severe disease, and on iMonos and ncMonos in severe COVID-19. While the role of the extracellular metalloprotease inducer CD147 that was discussed to be a viral spike protein receptor remains to be determined, it is also a potential host factor in infection-mediated coagulation^49^. In the present study, elevated levels of CD147 on iMonos and ncMonos was a general feature of all patients, while increased levels of CD147 on pDCs, pre-DCs and pre-DC2s were specific to severe COVID-19. In this context it is interesting to note that DC progenitors can themselves be susceptible to viral insult^50^. It remains to be addressed what causes CD147 upregulation in COVID-19 and how the MNP system may participate in dysregulated coagulation, but it has previously been shown that CD147 support platelet-monocyte interactions promoting vascular inflammation^51^. In addition, CD147 is upregulated upon TNF and IFN-γ stimulation, an effect strongly potentiated by bacterial stimuli^52^. Of note, the two pulmonary embolism cases in our cohort occurred in severe COVID-19 patients suffering from superinfection with positive blood cultures.

In summary, we here provide a comprehensive mapping of the MNP landscape in COVID-19. Lineage restricted changes were observed with loss of the inhibitory receptor CD200R in cDC2s, including the progenitors, associated with severe disease and fatal outcome as well as expansion of immature Mo-MDSC-like monocytes in critically ill. These findings suggest dysregulation of myeloid development in severe COVID-19, involving both monocyte and DC lineages, and changes detected in circulation were mirrored in lung tissue. Unsupervised analysis revealed that MNPs, alone, could identify a cluster with non-survivors. Further research is warranted to understand if and how immature myeloid derived cells in COVID-19 contribute to perpetuation and silencing of the inflammatory storm, and how the altered MNP development affects monocyte and DC ability to contribute to viral clearance. The new knowledge gained from investigating MNP biology provides a framework that has the potential to influence vaccine development, where accurate MNP host response are needed, and improvement of treatment strategies aiming at controlling the proinflammatory storm without affecting the immune system’s ability to clear the virus.

## MATERIALS AND METHODS

### Patient and healthy control cohorts

COVID-19 patients were recruited at the Karolinska University Hospital Stockholm. All patients were SARS-CoV-2 PCR positive in nasopharynx or sputum, had clinical COVID-19 disease, and were sampled within 8 days from hospitalization. Moderate COVID-19 patients and an oxygen saturation (SO_2_) between 90-94% or had a 0.5-3 L/min oxygen requirement at screening. Severe COVID-19 patients were treated at the intensive care unit or a high dependency unit. Patients with an ongoing malignancy or ongoing immune suppressive medication prior to hospitalization were excluded from the study. The patients were further described by Sequential Organ Failure Assessment (SOFA) score ^53^ and the National Institute of Health (NIH) ordinal scale ^54^. The NIH ordinal scale scores are as follows: 1, not hospitalized, no limitations; 2, not hospitalized, with limitations; 3, hospitalized, no active medical problems; 4, hospitalized not on oxygen but requiring ongoing medical care; 5, hospitalized, on oxygen; 6, hospitalized, on high flow oxygen or non-invasive mechanical ventilation; 7, hospitalized, on mechanical ventilation or ECMO; 8, death. In addition, a group of healthy volunteers matched for age and sex, no history of COVID-19 disease, no signs or symptoms of ongoing COVID-19, and SARS-CoV-2 IgG seronegative, was recruited in parallel as a control group. The research was approved by the Swedish Ethical Review Authority.

### Serology

SARS-CoV-2 IgG titers were measured at the day of sampling in serum in all the patients and controls according to the routinely used clinical pipeline at the Clinical Microbiology Department, Karolinska University Hospital; and finally interpreted by a clinical microbiologist as positive or negative, according to clinical practice.

### Viral titers

Viral load was assessed at the day of sampling as serum SARS-CoV-2 PCR positivity, examined in serum from all the patients following the pipeline at the Clinical Microbiology Department, Karolinska University Hospital. Briefly, two independent tests were used and finally interpreted by a clinical microbiologist as positive or negative, according to the clinical routine.

### Trucount staining

In order to determine the absolute leucocyte numbers, trucount staining was performed. Briefly, 50 μl EDTA blood and 20 μl of antibody mix (6-color TBNK Reagent and CD123 BUV395 from BD, CD15 PB, CD193 BV605 and HLA-DR BV785 from Biolegend and CD14 PE-Cy5 from eBioscience) were added in BD trucount tubes and incubated for 15 min at room temperature in the dark. The samples were then fixed using 430 μl 1X BD FACS lysing solution (BD Biosciences).

### Sample processing and flow cytometry

PBMCs were isolated from patient and control blood samples after Ficoll separation using Lymphoprep (Stemcell Technologies) and FACS staining was performed on freshly isolated cells in three independent experiments during three subsequent weeks to minimize batch effects. To assess batch related effects, internal control of frozen healthy PBMCs were used in all experiments and revealed minimal variability. Briefly, PBMCs were resuspended in PBS containing 2% FCS and 2 mM EDTA and a mixture of antibodies, supplemented with BD Horizon Brilliant Stain Buffer Plus (BD Biosciences) at 1:5 and FcR Blocking Reagent (Miltenyi Biotec) at 1:25 and were incubated for 30 min. Next, cells were washed twice and fixed in PBS containing 1% PFA for 2 hours in order to inactivate the virus. Cells were acquired on a FACSymphony A5 instrument (BD Biosciences), equipped with UV (355nm), violet (405 nm), blue (488 nm), yellow/green (561 nm), and red laser (637 nm). Details of antibodies used as well as information on filters for cytometer configuration are provided in Table 2.

### Flow cytometry analysis

Acquired data was analyzed using DIVA (BD Biosciences), FlowJo v.10.5.3 (BD Biosciences) and Prism v.8.0.2 (GraphPad Software Inc.). Absolute numbers of MNPs were determined relating to CD14+ cells in the trucount staining. Algorithms used for dimensionality reduction were UMAP (Becht et al., Nature Biotechnology, 2018; https://github.com/lmcinnes/umap) and Phenograph (Levine et al., Cell, 2015; https://github.com/JinmiaoChenLab/Rphenograph).

### Soluble factor analysis

Serum levels of soluble factors were analyzed at the day of the sampling using proximity extension assay, based on real-time PCR quantification of pair-wise binding of oligonucleotide-labeled target antibodies (Olink Proteomics, Uppsala, Sweden; sensitivity, specificity, dynamic range, repeatability, reproducibility and scalability validation documents are available at www.olink.com/downloads).

### Single cell analysis

Seurat version 3 was used to re-analyze single cell data. In brief, scRNA-seq from previously published report^22^ was used and 10X Genomics filtered_feature_bc_matrix files were acquired from Gene Expression Omnibus (GEO; accession number GSE145926). QC such as gene number between 200 and 6000, UMI count above 1000 and mitochondrial gene percentage below 0.1 were applied, and ‘LogNormalize’ method in Seurat V3 was used to normalize the data matrix followed by PCA using top 2000 most variable genes. Top 50 principal components were used to visualize the cells in UMAP, and ‘bimod’ test was used to detect differentially expressed genes. First, we selected myeloid DC clusters 22, 25, 27 (CLEC9A, CADM1, CD1c, FCER1A), pDC cluster 28 (LILRA4, IL3RA), and other myeloid clusters 0, 1, 2, 3, 4, 5, 6, 9, 10, 11, 12, 13, 21, 23, 29 (CD14, CD68), after exclusion of B cells, plasma cells, epithelium, NK cells, and T cells (Supp. Fig. 1A-F). After re-running total myeloid cells through the dimensionality reduction pipeline (Supp. Fig. 1G-K), neutrophils were excluded (Supp. Fig. 1F, G). To identify DCs, DC-containing clusters 11, 27, and 29 from the myeloid cell UMAP were re-clustered (Supp. 1L-P). As cluster 4 contained a comparable number of cells in severe and moderate cohorts, and cluster 7 in severe and healthy, the DEGs in those clusters were calculated between the severe patients and the respective cohort and subjected to pathway analysis. Ingenuity Pathway Analysis (IPA) was performed to study pathways (Content version: 51963813; Release Date: 2020-03-11; Ingenuity Systems). Predicted upregulation and downregulation of pathways was based on Z-score, where positive score implied upregulation and negative score implied downregulation.

### PCA and correlation plots

PCA plots were created using ‘prcomp’ function and ‘ggbiplot’ package in R and correlation mapping was performed using ‘corrplot’ package in R. The color of the circles indicated positive (red) and negative (blue) correlations, color intensity represented correlation strength as measured by the Pearson’s correlation coefficient. The correlation matrix was reordered using “hclust” for hierarchical clustering order. Significance tests were performed to produce p-values and confidence intervals for each pair of input features.

### Statistical analysis

Differences between two groups were evaluated using Mann-Whitney test, and Kruskal-Wallis test with Dunn’s multiple comparisons test were used to evaluate differences among the three groups in all the analysis. Friedman test was used in Fig. 2E to evaluate differences among paired variables. Spearman test was used for correlations of non-normally distributed data and Pearson test was used for normally distributed data. Significance for pathways analyses in Fig. 3E, 3H was defined by the IPA software (Ingenuity Systems). Significance level: *p<0.05, **p<0.01, ***p<0.001, ****p<0.0001.

## Data Availability

Curated flow cytometry data is available for exploration via the Karolinska COVID-19 Immune Atlas (homepage currently under construction).

## ACKNOWLEDGEMENTS

We thank the patients for their generous participation. We thank nurses, research nurses, physiotherapists, nurse assistants, dieticians, occupational therapists, physicians, psychologists, counselors, clinical laboratory analysis personnel and all other medical care personnel in Sweden and worldwide for their contributions during the COVID-19 pandemic. The study was supported by Knut and Alice Wallenberg Foundation, Nordstjernan AB, the Swedish Children’s Cancer Foundation, Swedish Cancer Foundation, Mary Béves foundation, and Erik och Edith Fernströms foundation.

## AUTHOR CONTRIBUTIONS

Conceptualization, E.K., M.S., N.K.B, H.G.L, and Karolinska COVID-19 Study Group; Methodology, E.K.; Formal analysis, E.K, L.H., I.H., I.H.M.; Investigation, E.K., L.H., A.P., M.L., M.D., M.A., J.K., J.R.M., P.C., K.S., E.F., O.R.; Recourses: E.F., L.I.E., O.R., S.A., K.S., S.B., A.N.T., F.G.; Writing – Original Draft, E.K.; Writing – Review and Editing, E.K., M.S., J-I.H., N.K.B., F.G.; Funding Acquisition, H.G.L, J-I.H., and E.K., Supervision, E.K., M.S.

## SUPPLEMENTARY MATERIAL AND METHODS

### Detailed cohort description

To address general features of the clinical biomarker pattern of COVID-19 we investigated parameters measured in the clinic as a part of routine monitoring, including inflammatory, hematological, biochemical and coagulation related parameters, and compared them between the two cohorts of COVID-19 patients, reference values indicated in grey (Figure 6A). As expected, almost all patients had elevated levels of almost all inflammatory parameters previously associated with COVID-19, such as CRP, ferritin, as well as the cytokine levels (IL-10, IL-6, IL-1β, TNF), and differences in IL-6 levels was detected between the two study cohorts with higher levels in the critically ill patients. Coagulation parameters were also affected, and higher levels of D-dimer were detected in the severe patients, who also had elevated levels of fibrinogen (85%, 11/13). Biochemical status revealed higher levels of lactate dehydrogenase (LDH) and myoglobin in critically ill patients and hypoalbuminemia in all, with lower levels in severe disease. From the hematological perspective, severe patients had higher levels of WBC, higher neutrophil counts, lower lymphocytes and subsequently higher neutrophil to lymphocyte ratio, with no striking differences in platelets or monocytes (Figure 6A). Radiologically, all patients examined showed bilateral infiltrates on chest x-ray; therapeutically, the majority received anticoagulants (93%, 25/27) and more than half broad-spectrum antibiotics (56%, 15/27). From a HLH perspective, all patients had fever at admission (100%, 27/27) and elevated ferritin levels (100%, 26/26, data not available for 1 patient), but no suspicion from the hematological parameters available, and in line rather elevated than low fibrinogen levels. Four patients in the severe group had superinfections (24% of severe patients, 4/17), in two of whom pulmonary embolism was detected, representing all detected cases of pulmonary embolism. To further address clinical laboratory status in relation to each other clinical parameters and outcome, integrative correlation mapping was performed (Figure 6B).

This showed cytokines (IL-10, TNF, IL-6), LDH, neutrophil and D-dimer levels clustering together with fatal outcome, peak oxygen need and viremia. Of note, levels of monocytes showed a positive correlation with body mass index (BMI), a known COVID-19 risk factor, and monocytes were the only clinically available cell type correlating with days from symptom debut, indicating the myeloid mononuclear cell importance in disease development timeline. We also investigated symptom panorama at admission, and found that in addition to fever, all patients had dyspnea (100%, 27/27), most had cough (85%, 23/27), few had GI related symptoms (15%, 4/27), while body ache showed higher variation (44%, 12/27), and correlated with thromboembolism, days since symptom debut and, intriguingly, levels of monocytes (Figure 6B). Importantly, a similar pattern was observed when integrative correlation mapping was performed including clinical parameters available 24 hours within MNP profiling (Supplementary Figure 4A). Further details on clinical and laboratory findings are presented in Supplementary Table 1 and 2.

**SUPPLEMENTARY TABLE 1.** Clinical characteristics.

|  | **Moderate** | **Severe** |
| --- | --- | --- |
| Group size, n | 10 | 17 |
| **Risk factors** |  |  |
| Age, median (range) | 56.5 (18-76) | 58 (40-78) |
| Sex, n (female/male) | 3/7 | 3/14 |
| BMI, median (range) | 27.63 (23- 35.06) | 29 (23-55) |
| Smoking, n (%) -current | 1 (10) | 2 (12) |
| - prior | 2 (20) | 5 (29) |
| - non-smoker | 2 (20) | 9 (53) |
| - unknown | 5 (50) | 1 (6) |
| **Comorbidities, n (%)** |  |  |
| None | 4 (40) | 5 (29) |
| Hypertension | 1 (10) | 6 (35) |
| Diabetes | 3 (30) | 5 (29) |
| Chronic heart disease | 1 (10) | 2 (12) |
| Asthma | 1 (10) | 2 (12) |
| Other | Obstructive sleep apnea syndrome (OSAS) | Renal failure, OSAS, Latent Tuberculosis, Chronic hepatitis B infection |
| **Peak supportive oxygen therapy during hospitalization, n (%)** |  |  |
| None | 2 (20) | 0 (0) |
| Low flow 0.5-10L/min | 6 (60) | 0 (0) |
| Low flow 10–15 L/min | 1 (10) | 3 (18) |
| High Flow | 1 (10) | 1 (6) |
| Ventilator | 0 (0) | 12 (71) |
| ECMO | 0 (0) | 1 (6) |
| **Lung function/severity scores at sampling** |  |  |
| PaO<sub>2</sub>/FiO<sub>2</sub> (PF) ratio mmHg, mean (range) | 369 (312-523) | 139 (52-285) |
| SOFA respiration *, median (range) | 1 (0-1) | 3 (2-4) |
| SOFA score, median (range) | 1 (0-2) | 6 (2-12) |
| NIH Ordinal Scale, median (interquartile range) | 5 (5-5) | 7 (6-7) |
| **Outcome** |  |  |
| Discharged, n (%) | 10 (100) | 12 (71) |
| Deceased, n (%) | 0 (0) | 4 (24) |
| Ongoing treatment, n (%) | 0 (0) | 1<sup>#</sup> (6) |

\* Part in SOFA score regarding respiratory system.
<sup>#</sup> Still in ECMO (26 June 2020).
BMI, body mass index; ECMO, extracorporeal membrane oxygenation; IL, interleukin; NA not available; TNF, tumor necrosis factor.

**SUPPLEMENTARY TABLE 2.** Individual patient characteristics.

| Patient | Severity group | Days from symptom debut to admission | Days from symptom debut to sampling | Blood culture +/- 5 days from sampling | Culture finding in lower resp. secr. +/- 5 days from sampling | Steroids* | Antibiotics* | COVID-19 therapy* |
| --- | --- | --- | --- | --- | --- | --- | --- | --- |
| Patient 1 | Moderate | 7 | 14 | 0 | 0 | No | Cefotaxim | 0 |
| Patient 2 | Moderate | 7 | 14 | 0 | 0 | Yes | Cefotaxim | 0 |
| Patient 3 | Moderate | 12 | 17 | 0 | 0 | No | 0 | 0 |
| Patient 4 | Moderate | 8 | 10 | 0 | 0 | No | 0 | 0 |
| Patient 5 | Moderate | 10 | 13 | 0 | 0 | No | 0 | 0 |
| Patient 6 | Moderate | 7 | 14 | 0 | 0 | No | 0 | 0 |
| Patient 7 | Moderate | 4 | 6 | 0 | 0 | No | 0 | 0 |
| Patient 8 | Moderate | 9 | 11 | 0 | 0 | No | 0 | 0 |
| Patient 9 | Moderate | 14 | 19 | 0 | 0 | Yes | Cefotaxim | 0 |
| Patient 10 | Moderate | 9 | 11 | 0 | 0 | No | 0 | 0 |
| Patient 11 | Severe | 7 | 14 | 0 | 0 | No | Cefotaxim | 0 |
| Patient 12 | Severe | 7 | 14 | 0 | *S. dysgalactiae*, *S. aureus* | No | 0 | 0 |
| Patient 13 | Severe | 9 | 12 | *S. aureus* | *S. aureus* | Yes | 0 | 0 |
| Patient 14 | Severe | 3 | 11 | *S. milleri* | *S. aureus*, *K. pneumoniae*, *A. fumigatus*, *C. albicans* | Yes | Cefotaxim | 0 |
| Patient 15 | Severe | 5 | 12 | 0 | 0 | Yes | Piperacillin/Tazobactam | 0 |
| Patient 16 | Severe | 14 | 19 | 0 | 0 | No | Cefotaxim | 0 |
| Patient 17 | Severe | 12 | 16 | 0 | 0 | Yes | Cefotaxim | 0 |
| Patient 18 | Severe | 13 | 20 | 0 | 0 | Yes | Cefotaxim | 0 |
| Patient 19 | Severe | 4 | 5 | 0 | 0 | No | 0 | 0 |
| Patient 20 | Severe | 7 | 13 | *S. aureus* | *S. aureus*, *H. influenzae* | No | 0 | Remdesivir |
| Patient 21 | Severe | 9 | 15 | 0 | 0 | Yes | Cefotaxim | Tocilizumab |
| Patient 22 | Severe | 6 | 10 | 0 | 0 | Yes | 0 | 0 |
| Patient 23 | Severe | 11 | 15 | 0 | 0 | Yes | Cefotaxim | 0 |
| Patient 24 | Severe | 12 | 17 | 0 | 0 | Yes | Cefotaxim | Anakinra |
| Patient 25 | Severe | 4 | 24 | 0 | *E. coli* | Yes | Cefotaxim,<br>Piperacillin/Tazobactam | 0 |
| Patient 26 | Severe | 14 | 17 | 0 | *C. albicans* | Yes | Cefotaxim | 0 |
| Patient 27 | Severe | 9 | 14 | *S. aureus*, *E. faecalis* | *S. aureus* | Yes | Cefotaxim | 0 |

\* Started at hospital prior to sampling

**SUPPLEMENTARY TABLE 3.**
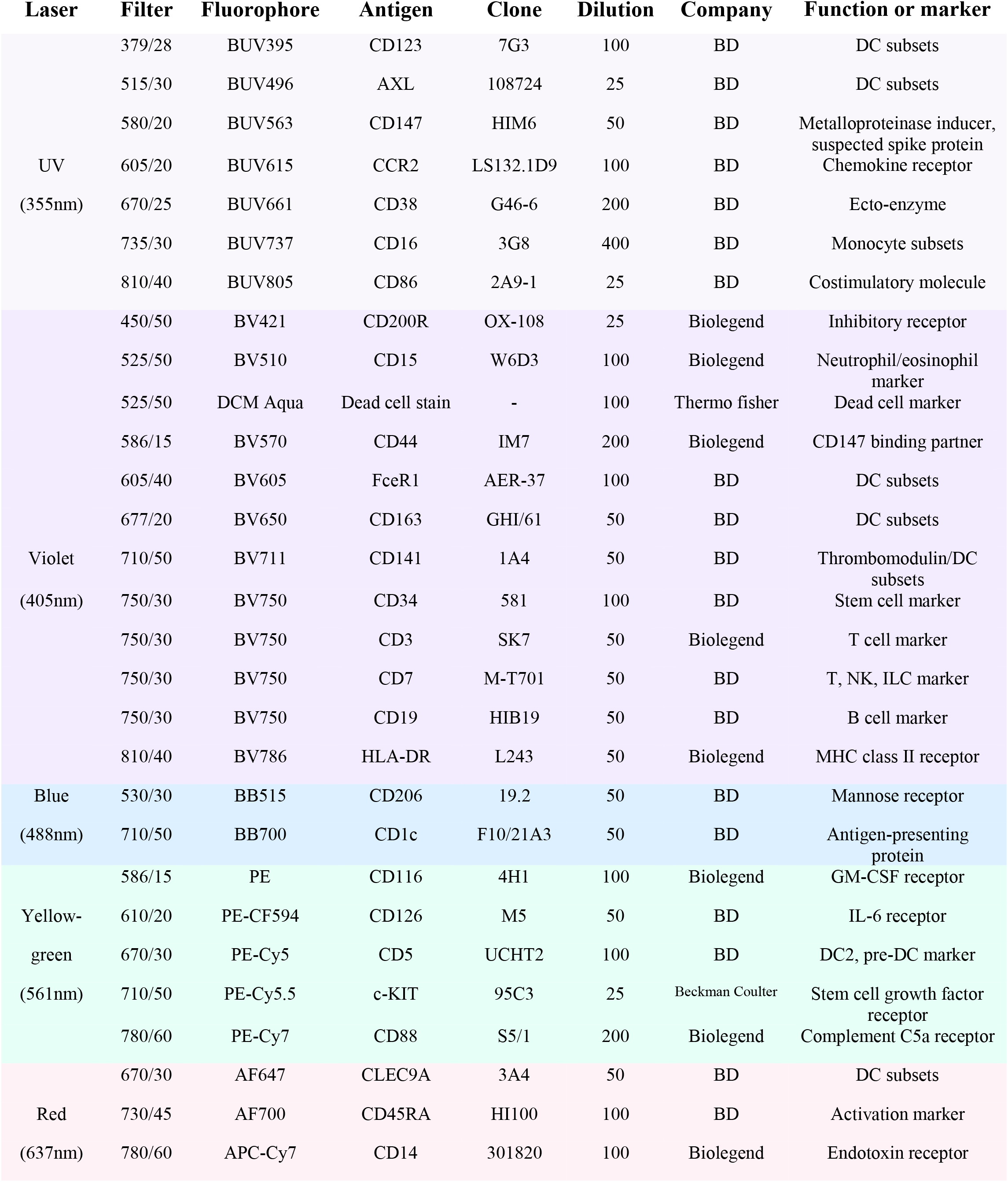
Antibodies.

**SUPPLEMENTARY FIGURES AND FIGURE LEGENDS**

**SUPPLEMENTARY FIGURE 1:**
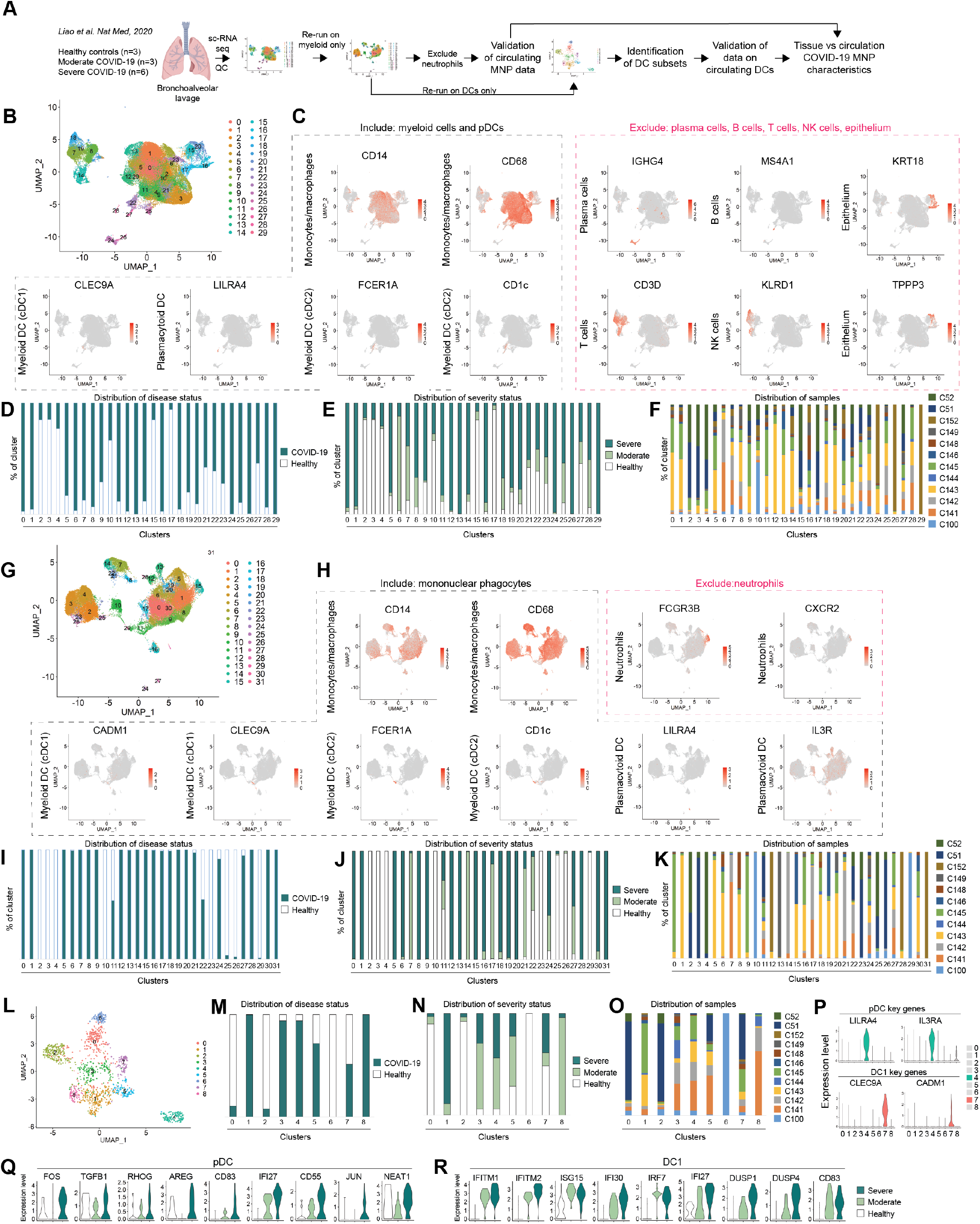
sc-RNAseq re-analysis (related to Fig. 3, Fig. 4, and Methods section “Single cell analysis”) (A) Analytic pipeline of sc-RNAseq data. (B) After quality control, scRNA-seq data reanalyzed using the Seurat version 3 (V3). (C) Expression plots of key markers used to discriminate between cells to be included and excluded in subsequent analysis. (D) Distribution of cells from COVID-19 patients and healthy controls among clusters generated in the whole data set. (E) Distribution of cells from moderate and severe COVID-19 patients and healthy controls among clusters generated in the whole data set. (F)Distribution of cells from each sample among clusters generated in the whole data set. (G) Selected myeloid cell clusters re-analyzed. (H) Expression plots of key markers used to discriminate between MNP and neutrophil clusters. (I) Distribution of cells from COVID-19 patients and healthy controls in MNP clusters. (J) Distribution of cells from moderate and severe COVID-19 patients and healthy controls in MNP clusters. (K) Distribution of cells from each sample in MNP clusters. (L) Clusters containing DCs selected and reanalyzed. (M) Distribution of cells from COVID-19 patients and healthy controls in DC clusters. (N) Distribution of cells from moderate and severe COVID-19 patients and healthy controls in DC clusters. (O) Distribution of cells from each sample in DC clusters. (P) Key genes of pDC and DC1 in cluster 4 (green) and 7 (red), respectively. (Q) Genes in pDCs differentially expressed between severe and moderate COVID-19 patients, presented in the three cohorts. (R) Genes in DC1s differentially expressed between severe COVID-19 and healthy controls, presented in the three cohorts. Statistical evaluation using ‘bimod’ test for differentially expressed genes.

**SUPPLEMENTARY FIGURE 2:**
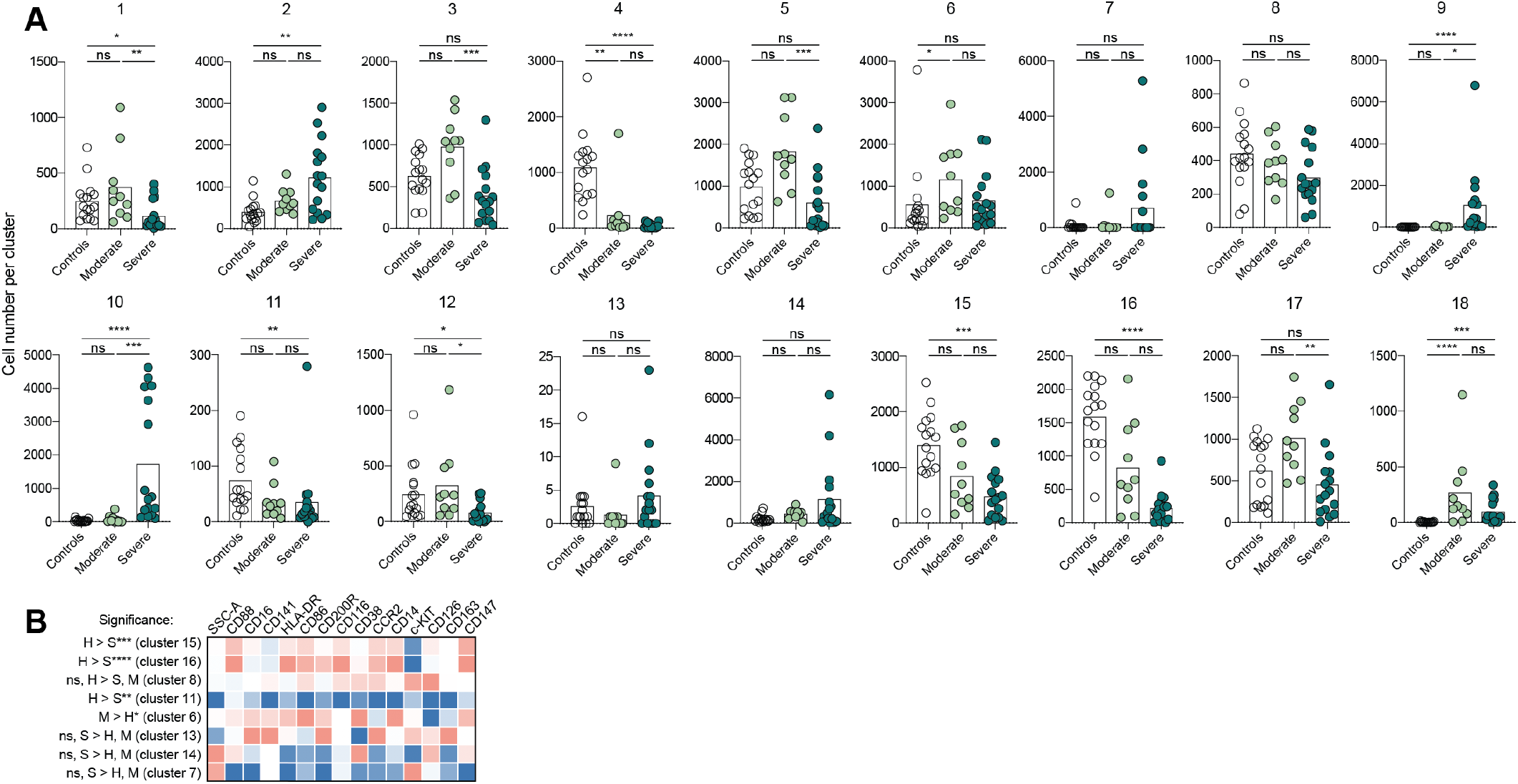
Monocyte clusters (related to Fig. 4) (A) Differences in cell numbers per Phenograph cluster 1-18 among the three cohorts. (B) Expression of markers in the remaining Phenograph clusters among the three cohorts. Statistical evaluation using Kruskal-Wallis test and Dunn’s multiple comparisons test for comparison between the three cohorts. Significance level: *p<0.05, **p<0.01, ***p<0.001, ****p<0.0001

**SUPPLEMENTARY FIGURE 3:**
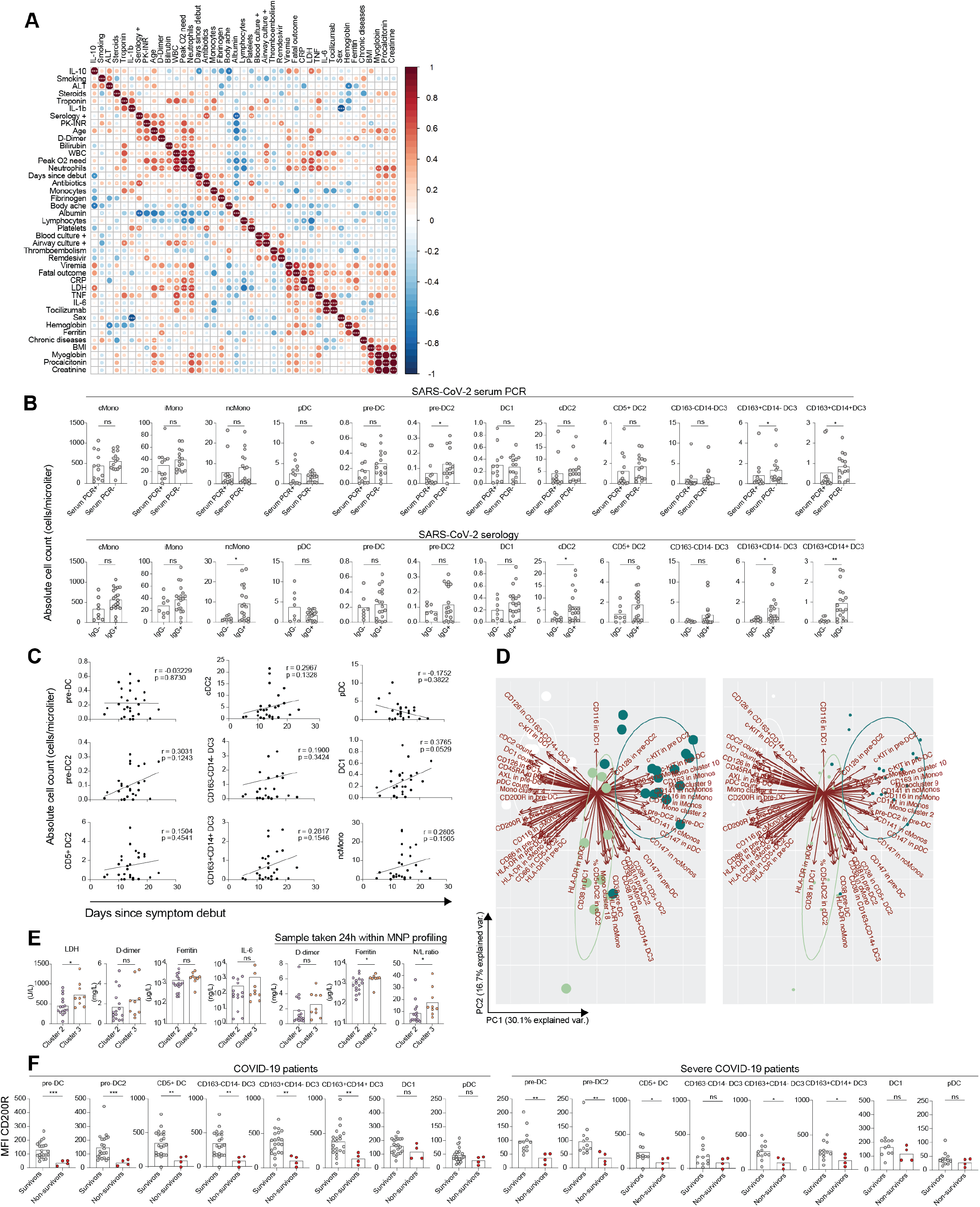
Clinical hyperinflammation, MNP landscape, and COVID-19 outcome (related to Fig. 5) (A) Integrated correlation clustering of clinical parameters with CRP, procalcitonin, hemoglobin, WBC, lymphocytes, neutrophils, ferritin, troponin, D-dimer, PK-INR, fibrinogen, creatinine, bilirubin, and albumin taken within 24 hours from the MNP profiling; the color of the circles indicated positive (red) and negative (blue) correlations, color intensity represented correlation strength as measured by the Pearson’s correlation coefficient. (B) Differences in absolute numbers of MNP populations between SARS-CoV-2 PCR+ and SARS-CoV-2 PCR- patients, and between SARS-CoV-2 IgG- and SARS-CoV-2 IgG+ patients. (C) Correlation of days since symptom debut until the sample collection and absolute numbers of cells within MNP populations. (D) Principal component analysis of 108 MNP parameters in the three cohorts, parameters contributing most to the separation highlighted; each dot represents controls (white), moderate COVID-19 (light green), and severe COVID-19 (dark green) patient; to better visualize marker names, a plot with smaller dots is provided (right). (E) Differences in LD, D-dimer, Ferritin, neutrophils/lymphocyte ratio at peak or 24h within MNP profiling (lower row) between patient cluster 2 and 3. (F) Differences in CD200R MFI between survivors and non-survivors in all patients (left) and in severe patients only (right). Statistical evaluation using Mann-Whitney for comparison between the two groups, Spearman test for correlations of non-normally distributed data and Pearson test for correlations of normally distributed data. Significance level: *p<0.05, **p<0.01, ***p<0.001, ****p<0.0001.

